# Increased neurotoxicity due to activated immune-inflammatory and nitro-oxidative stress pathways in patients with suicide attempts: a systematic review and meta-analysis

**DOI:** 10.1101/2021.04.16.21255605

**Authors:** Asara Vasupanrajit, Ketsupa Jirakarn, Chavit Tunvirachaisakul, Michael Maes

## Abstract

**Background:** Suicide attempts (SA) frequently occur in patients with mood disorders and schizophrenia, which are both accompanied by activated immune-inflammatory and nitro-oxidative (IO&NS) pathways.

**Methods:** We searched PubMed, Google Scholar, and Web of Science, for articles published from inception until February 1, 2021. We included studies that compared blood biomarkers in psychiatric patients with (SA+) and without SA (SA-) and heathy controls and we combined different IO&NS biomarkers into immune, inflammatory, and neurotoxic profiles and used meta-analysis (random-effect model with restricted maximum-likelihood) to delineate effect sizes with 95% confidence interval (CI).

**Findings:** Our search included 51 studies comprising 4.945 SA+ patients and 24.148 controls. We stratified the control group into healthy controls and SA-patients. SA+ patients showed significantly (p<0.001) increased immune activation (SMD: 1.044; CI: 0.599-1.489), inflammation (SMD: 1.109; CI: 0.505, 1.714), neurotoxicity (SMD: 0.879; CI: 0.465, 1.293), and lowered neuroprotection (SMD: 0.648; CI: 0.354, 0.941) as compared with healthy controls. When compared with SA-patients, those with SA+ showed significant (p<0.001) immune activation (SMD: 0.290; CI: 0.183, 0.397), inflammation (SMD: 0.311; CI: 0.191, 0.432), and neurotoxicity (SMD: 0.315; CI: 0.198, 0.432), and lowered neuroprotection (SMD: 0.341; CI: 0.167, 0.515). Patients with current, but not lifetime, SA showed significant (p<0.001) levels of inflammation and neurotoxicity as compared with controls.

**Conclusions:** Patients with immune activation are at a higher risk of SA which may be explained by increased neurotoxicity due to inflammation and nitro-oxidative stress. This meta-analysis discovered new biomarkers of SA and therapeutic targets to treat individuals with SA.

## Introduction

Suicide is a mental health problem globally causing about 800,000 deaths each year.^1^ More than half of the suicide deaths (52.1%) are under the age of 45 years and the prevalence in males is 1.8 times higher than is females.^1^ The Columbia Classification Algorithm of Suicide Assessment^2^ describes the main types of suicidal behaviors including completed suicide (CS), suicide attempts (SA), and suicidal ideation (SI). CS is a fatal self-injurious behavior, whereas SA and SI are non-fatal suicidal behaviors. SA can be classified into subgroups comprising non-violent, violent, and recurrent suicide attempts^3^. Non-violent SA is a method with a lower risk of death (including drug overdoses or superficial wrist cutting), whereas violent SA yields a higher risk to life (e.g., jumping from the high place, hanging, drowning, etc.).

Previous meta-analysis reported major risk factors of suicidal behaviors including internalizing psychopathology (e.g., mood disorders, hopelessness, etc.), demographics (e.g., age, sex, education, ethnicity, socioeconomic status, etc.), externalizing psychopathology (e.g., impulsivity, substance abuse, etc.), prior suicidal behaviors, and psychosocial factors (e.g., history of abuse or trauma, stressful life events, etc.).^4^ A meta-analysis of 3.275 CS cases indicates that there is a strong association between CS and affective disorders, especially with depressive episodes and psychotic disorders.^5^

There is robust evidence that major depressive disorder (MDD), bipolar disorder (BD), and schizophrenia are accompanied by activation of the immune-inflammatory response system (IRS) and activation of the compensatory immune-regulatory reflex system (CIRS), which downregulates an overzealous IRS.^6, 7^ IRS biomarkers include interleukin (IL)-1β^8, 9^, IL-6^8, 10^, IL-8^11^; interferon gamma (IFN-γ)^12, 13^; tumor necrosis factor alpha (TNF-α)^9, 10^; neutrophil-to-lymphocyte ratio (NLR)^14^; chemokines^15^; etc.), and CIRS biomarkers, e.g. IL-4^12^, transforming growth factor beta (TGF-β)^16, 17^; and soluble IL-2 receptor (sIL-2R)^18^. These CIRS products are released during an IRS response and exert a negative feedback on diverse aspects of the IRS and, therefore, increased levels in those CIRS biomarkers indicate IRS activation, but at the same time indicate increased immune-regulatory activities.^7^

Other immune biomarkers of mood disorders and schizophrenia are the presence of an acute phase or inflammatory response characterized by an increased production of positive acute phase proteins including C-reactive protein (CRP) and fibrinogen, and a downregulation of negative acute phase proteins such as albumin.^19, 20^ The immune-inflammatory response in mood disorders is also accompanied by induction of the tryptophan catabolite (TRYCAT) pathway.^21^ Both the acute phase response (APR) and production of TRYCATs are induced by pro-inflammatory cytokines including IL-1β, TNF-α, and IL-6.^22, 23^

Immune activation is frequently associated with activation of oxidative and nitrosative stress (O&NS) pathways and with lowered levels of key antioxidants such as high-density lipoprotein cholesterol (HDL-c) and vitamins including vitamin D.^24, 25^ There is now evidence that both O&NS and lowered antioxidant levels play a key role in the pathophysiology of affective disorders and schizophrenia.^26, 27^ In those disorders, increased nitro-oxidative stress in indicated by elevated levels of lipid hydroperoxides (LOOH) and malondialdehyde (both indicating lipid peroxidation), advanced oxidation protein products (indicating increased oxidation of proteins) and nitric oxide (NO) metabolites (NOx).^27, 28^ Moreover, both disorders are accompanied by lowered levels of key antioxidants including HDL-c, vitamin D, and albumin.^29^

The neurotoxicity theories of mood disorders and schizophrenia consider that the neurotoxic effects of immune and O&NS (IO&NS) pathways cause dysfunctions in gray and white matter functional plasticity (including axon sprouting, synaptogenesis, glial changes, neurogenesis, and myelin formation) thereby explaining the onset of affective disorders and schizophrenia.^30–32^

The first inkling that suicidal behaviors may be associated with immune functions and oxidative stress were published in the1990s.^18, 33, 34^ Since then, reports showed that increased levels of cytokines^35, 36^, C-reactive protein (CRP)^37^, fibrinogen^38^, and TRYCATs^39^, an increased erythrocyte sedimentation rate (ESR)^38^, and lowered albumin^40^ levels are associated with suicidal behaviors. Nevertheless, there are also contradictory results in suicide research. For example, a study^36^ showed lower IL-2 and IL-4, but higher TGF-β plasma levels in severe SI and SA patients as compared with healthy controls, whereas another study^35^ showed no significant associations between IL-4 and TGF-β plasma levels in patients with SA (SA+) as compared with patients without SA (SA-). Moreover, evidence suggests that CRP levels are significantly increased in depressive patients with suicidal behaviors,^37^ whereas other findings showed no differences in CRP levels between SA+ patients with acute depressive disorder and those without SA^8^. There are emerging data that suicidal behaviors are associated with activated O&NS pathways, including increased levels of AOPP, LOOH, and MDA and NOx.^38, 41^ A lifetime history of SA is also accompanied by lowered levels of plasma total radical-trapping antioxidant parameter (TRAP)^38^ and vitamin D^42^, both indicating lowered antioxidant defenses. On the other hand, results on HDL-c, another strong antioxidant, are contradictory with higher and lowered concentrations being reported in people with SA.^43^

Finally, a number of other immune-inflammatory and O&NS (IO&NS) biomarkers may be associated with suicidal behaviors. For example, lowered levels of one of the omega-3 polyunsaturated fatty acids (PUFAs), namely eicosapentaenoic acid (EPA), is significantly associated with major depression^44^ and with SA^45^. EPA has strong anti-inflammatory, antioxidant and neuroprotective properties and, therefore, lowered EPA levels predispose toward exaggerated IO&NS responses.^23^ A meta-analysis study showed decreased plasma levels of brain-derived neurotrophic factor (BDNF), a neurotrophic factor with antioxidant properties^46^, in subjects with SA.^47^ **Figure 1** and **Electronic Supplementary File (ESF), Table 1** show the IO&NS biomarkers that are have been described in patients with suicidal behaviors.

**Figure 1.**
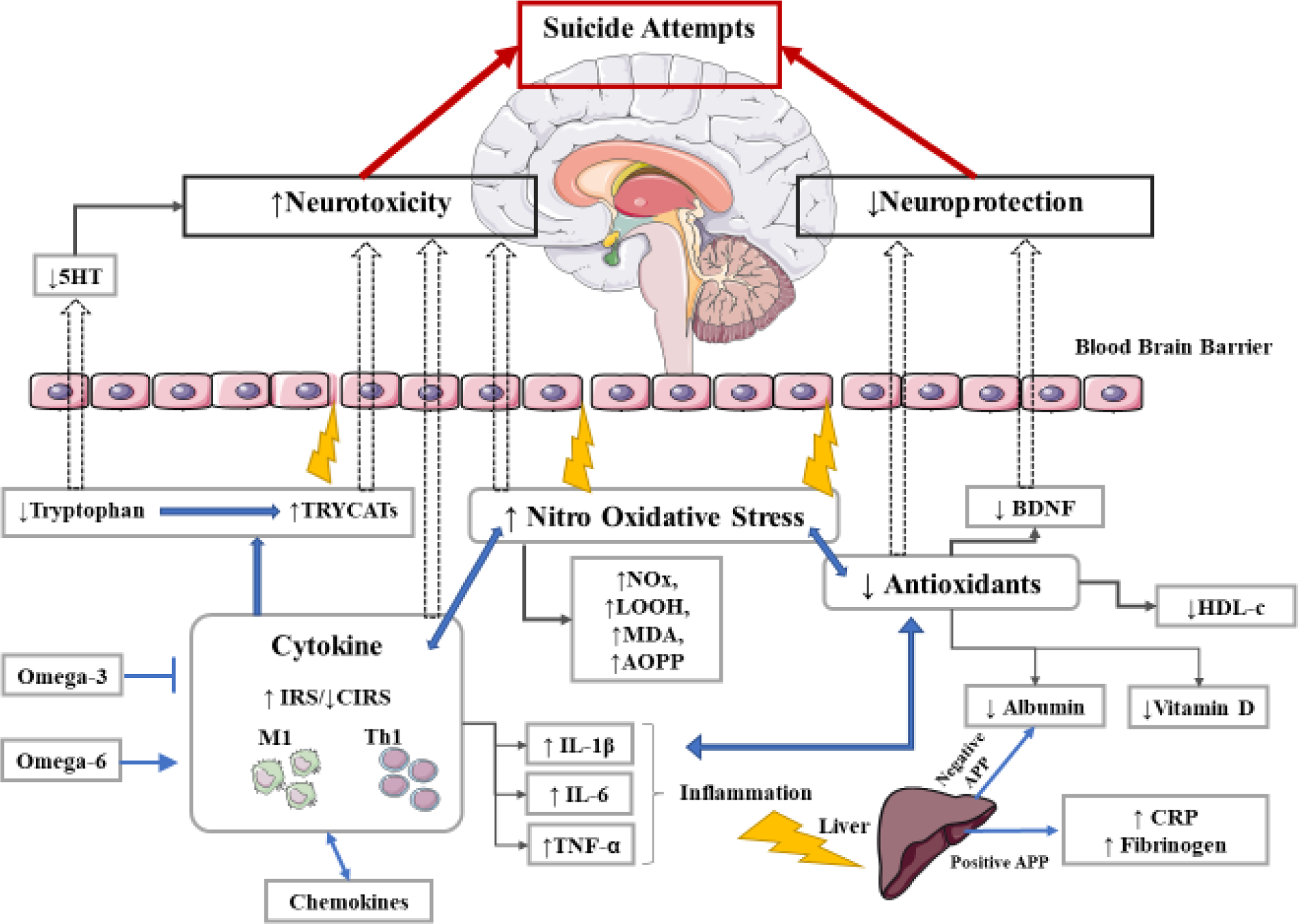
The pathophysiology of suicide attempts (SA) as characterized by different immune and nitro-oxidative stress-related biomarkers. Immune activation in patients with suicide attempts is indicated by signs of an activated immune-inflammatory response system including activated M1 macrophage and T helper (Th)-1 phenotypes, with increased production of pro-inflammatory cytokines including interleukin (IL)-6, IL-1β, and tumor necrosis factor alpha (TNF-α). The latter explain the presence of an acute phase response with increased production of positive acute phase proteins (APPs), including C-reactive protein (CRP) and fibrinogen, and negative APPs (including albumin). The production of cytokines is modulated by ω6 (stimulated) and ω3 (attenuated) polyunsaturated fatty acids. Increased M1 macrophage and Th-1 cytokines induce the catabolism of tryptophan into tryptophan catabolites (TRYCATs) yielding lowered tryptophan and 5-HT, and increased levels of neurotoxic and depressogenic TRYCATs. Immune activation is also accompanied by increased nitro-oxidative stress as indicated by higher nitric oxide metabolites (NOx), lipid hydroperoxides (LOOH), malondiadehyde (MDA), and advanced oxidation protein products (AOPP) and lowered antioxidant levels including high-density lipoprotein (HDL) cholesterol. Lowered levels of brain-derived-neurotrophic factor (BDNF) contribute to lowered antioxidant and neurotrophic protection. Increased levels of M1 and Th-1 cytokines, neurotoxic TRYCATs, and nitro-oxidative stress may lead to breakdown of the blood brain barrier (BBB) and thus increased entry of neurotoxic products and activated immune cells, probably leading to neuro-inflammation. Increased neurotoxicity and lowered antioxidant neuroprotection may induce changes in grey and white matter functional plasticity thereby causing suicidal behaviors.

**Table 1.** Number of patients with suicidal attempts (SA+) and controls in the different meta-analyses and side of standardized mean difference (SMD) and the 95% confidence intervals with respect to zero SMD.

| Outcome profiles | n studies | Side of 95% confidence intervals |  |  |  | SA+ patients<br>number | Controls<br>numbers | Total number<br>of<br>participants |
| --- | --- | --- | --- | --- | --- | --- | --- | --- |
|  |  | < 0 | Overlap 0<br>and SMD < 0 | Overlap 0<br>and SMD > 0 | > 0 |  |  |  |
| IO&NS | 51 | 0 | 6 | 20 | 25 | 4.945 | 24.148 | 29.093 |
| IRS | 33 | 0 | 3 | 12 | 18 | 3.669 | 19.863 | 23.532 |
| Inflammation | 22 | 0 | 3 | 6 | 13 | 3.401 | 19.160 | 22.560 |
| Neurotoxicity | 27 | 0 | 4 | 8 | 15 | 3.269 | 19.046 | 22.315 |
| ANTIOXPRO | 32 | 0 | 7 | 11 | 14 | 3.723 | 20.815 | 24.538 |

IO&NS: immune-inflammatory and oxidative and nitrosative stress
IRS: immune-inflammatory response system
ANTIOXPRO: protection via antioxidants and neurotrophic products.

Nevertheless, at present, there are no systematic reviews or meta-analyses, which examined the IO&NS, inflammatory, neurotoxic and antioxidant/neuroprotective (ANTIOXPRO) profiles in people with suicidal attempts. Hence, the current study aimed to review and meta-analyze the associations between current and a lifetime history of SA and IO&NS, inflammation, neurotoxic and ANTIOXPRO biomarker profiles. Based on previous knowledge, we hypothesized that IO&NS, inflammation, and neurotoxicity profiles are activated in SA+ patients and that the ANTIOXPRO profile may be lowered in SA+ patients as compared with SA-patients and healthy controls.

## Materials and methods

The methodological approach was accomplished following to the Preferred Reporting Items for Systematic Reviews and Meta-Analyses (PRISMA) guidelines, as well as the guidelines of the Cohrane Handbook for Systematic Reviews and Interventions^48^, and the Meta-Analyses of Observational Studies in Epidemiology (MOOSE)^49^. No public or patient representatives were directly or indirectly involved in the draft or process of this study. This meta-analysis combines different biomarkers into composites reflecting IO&NS, IRS, inflammation, neurotoxicity and ANTIOXPRO activities and examines these profiles in SA+ patients, either lifetime or current, as compared with SA-patients and healthy controls.

### Search strategy

A systematic review was conducted by database inception, last search on February 1, 2021. We used the electronic databases PubMed/MEDLINE, Google Scholar, and Web of Science, for searching articles. The search used the major terms of immune-inflammatory and O&NS biomarkers and suicide, include “inflamm*” OR immun*”, “cytokine”, “chemokine”, “IL-6”, “IL-1”, “interleukin”, “C-Reactive Protein”, “CRP”, “tumor necrosis factor”, “TNF”, “interferon”, “IFN”, “Transforming growth factor”, “TGF”, “Tryptophan”, “Oxidative stress”, “Antioxidants”, “Zinc”, “Vitamin”, “Albumin”, “Nitric oxide”, “Lipid hydroperoxides”, “Omega 3”, “Coenzyme Q10”, “DHA”, “suicid*”. Moreover, a manual search was conducted on the reference list of the studies included and previous meta-analysis studies.

### Eligibility criteria

This search first selected manuscripts written in English and published in peer-reviewed journals, after which we searched additional records in the reference lists of the studies and grey literature. We included observational case-control and cohort studies which examined associations between SA and IO&NS biomarkers including in serum or plasma, but not cerebrospinal fluid (CSF), urine, platelets or stimulated whole blood, in patient and controls of both sexes, and all ages and ethnicities. Inclusion criteria were: a) observational case-control or cohort studies, which examined psychiatric patients with SA, either lifetime or current, and compared SA+ patients with SA-patients and/or healthy controls; b) studies including biomarkers belonging to the IO&NS, inflammation, neurotoxicity and ANTIOXPRO profiles as listed in **ESF Table 1**; and c) studies including patients diagnosed with MDD, BD, mixed mood disorder episodes, dysthymia, adjustment disorder with depressed mood, anxiety disorders including panic disorder, substance-induced mood disorders, schizophrenia spectrum disorders, and borderline personality disorder as defined according to the diagnostic criteria of DSM or ICD-any version. We excluded a) studies reporting animal models, and translational and genetic research results; b) case reports, case series, and cohort studies without a control group; c) systematic reviews and meta-analysis; d) studies not focusing on the clinical groups as defined above; e) studies not reporting on the predefined IO&NS, inflammation, neurotoxicity or ANTIOXPRO biomarkers; f) studies which presented duplicated data; and g) studies that did not report mean with SD/SE values, as for example, geometric means, median values or mean/median values shown in graph format. Nevertheless, the authors of the latter studies were contacted and requested to provide means, SD, and number of cases and controls. When the authors did not respond to our request and showed median with either interquartile range (IQR) or minimum/maximum values, we estimated mean and SD values based on formulas provided by Wan et al.^50^ We recorded any reasons why studies were excluded from the analysis.

### Primary and secondary outcomes

The primary outcome was the pooled standardized mean difference (SMD) of the IO&NS profile in SA+ patients (either current or lifetime) versus SA- patients and healthy controls combined. In order to delineate which component of the IO&NS is more relevant to SA, we also examined subdomains of the IO&NS profile, namely IRS, inflammation, neurotoxicity, and ANTIOXPRO composites, which were the secondary outcomes.

### Screening and data extraction

The first author (AV) made a first screening of the studies and assessed eligibility by considering the titles and abstracts of the searched papers and collected the full text of the potentially eligible papers. Consequently, the first author of this study (AV) derived the key data into a predefined Excel spread sheet from the full-text versions. The second author (KJ) checked the extracted data once the first author (AV) completed all information. Furthermore, the last author (MM) was consulted in case of disagreement.

A predefined Excel spread sheet template was employed consisting of the key data from all eligible studies comprising the author’s name, publication year, IO&NS profile, biomarker type, mean concentration levels with SD and sample size, SA current or lifetime and type of controls either SA-patients or healthy volunteers. Moreover, we included the following information extracted from each study: study setting and design, socio-demographic data of the study groups (mean age, sex and ethnicity distribution, psychiatric disorders of participants, violent and non-violent SA defined according to Paykel and Rassaby^3^, assessment of SA, medium (plasma or serum), latitude of the study, and methodological quality scores of the study. We used a methodological quality score checklist, namely the immune cofounder’s scale (ICS)^51^, which was slightly modified by the last author. **ESF, Table 3** shows the quality and red point score checklists used in our meta-analysis. The first part aims to assess methodological quality of IO&NS studies, considering the more critical aspects including sample size, confounder control, specific time for sample collection, etc. A score is calculated to estimate the methodological quality which may vary from 0 to 10, with higher scores indicating better methodological quality. The second part includes a red-points scale which scores the severity of neglecting to control for critical confounders (see ESF, Table 3), which may induce considerable bias in IO&NS studies. Total scores may range from 0 to 26, with 0 indicating that all the confounder variables were considered and 26 when there was no control at all.

### Data analysis

A PRISMA-compliant systematic review and meta-analysis were conducted, and statistical analyses were performed using the CMA V3 software. This review and meta-analysis considered five outcome variables, namely IO&NS, IRS, inflammation, neurotoxicity and ANTIOXPRO profiles. **ESF, Table 1** shows the composition of the different outcome variables or summary effects examined here. A meta-analysis was performed whenever values of the outcome variables were available in three or more studies. When more than one biomarker reflecting one of the profiles was reported in the same study, a synthetic score was computed for each study in order to avoid dependent measurements, which would artificially narrow the confidence intervals. Thus, in the CMA, we used multiple outcome profiles accommodating any number of biomarkers based on the same participants and these synthetic scores were compared among SA+ patients *versus* SA-patients and healthy controls. The direction of all immune-inflammatory and neurotoxic variables was set as positive (thus increased levels favor SA+), whereas that of ANTIOXPRO biomarkers was set as negative (thus lowered levels favor SA+).

This study used the random-effects model with restricted maximum-likelihood for the study under the assumption that there were differences between population’s characteristics in the different studies. We estimated the SMD with 95% confidence intervals (CI). The SMD was used because it provides an unbiased effect size adjusted for small sample sizes and because the studies reported different methods to assay biomarkers. An alpha level of 0.05 indicated statistically significant results (two sided tests). SMD values around 0.2, 0.5, and 0.8 indicate small, moderate, and large effect sizes, respectively.^52^ Moreover, we also computed the prediction intervals.^53^

Heterogeneity among studies was evaluated using the Cochrane Q test, which probes homogeneity of the study-specific effect sizes, and the I^2^ metric, which reflects the variability (in %) due to heterogeneity rather than sampling error. According to the Cohrane Handbook for Systematic Reviews and interventions^48^ important ranges for interpretation are: I^2^ 0-40%: heterogeneity may not be important; 30-60%: may represent moderate heterogeneity; 50-90%: may represent substantial heterogeneity; and 75-100%: considerable heterogeneity. Some authors employ a I^2^ ≥50% threshold value to indicate significant hereogeneity.^54^ Tau (τ), which is the estimated standard deviation of the true mean differences across the studies included, and Tau^2^ which is its variance, are better indices of heterogeneity.^55^ In the case of heterogeneity especially when τ^2^ is imprecise, random-effects subgroup meta-analysis are performed considering the *a priori* defined patient subgroups (namely lifetime versus current SA), and control subgroups (SA-patients and healthy controls).

Potential sources of heterogeneity across studies were investigated when at least 10 studies reported data of the same profile, using either subgroup meta-analysis (with a minimum of three studies per sub-group) or random-effects meta-regression analyses (at least 10 studies).^56^ The group-by-analyses were checked using within- and between-group heterogeneity results. We used the study as well as the prespecified subgroups as the units of analysis. Thus, subgroup analysis compared the pooled effect size of IO&NS, IRS, inflammation, neurotoxicity and ANTIOXPRO profiles in a) SA+ versus SA-patients and healthy controls; and b) SA+ patients with a lifetime history or current SA versus SA-patients and healthy controls. In addition, we also examined potential sources of heterogeneity through group analysis and meta-regression using variables which were not defined a priori. These exploratory analyses considered the male to female ratio, ethnicity, country, latitude dichotomized using 40°, medium of biomarkers (serum, plasma), type of psychiatric disorders (MDD, affective disorders, other diagnoses, or mixed patient groups), suicide methods (violent and non-violent), exclusion of medical conditions (yes/no), and time of blood sample (morning vs. not reported) in group analysis; and the red point and quality scores and latitude in meta-regression analysis.

We also performed sensitivity analyses to evaluate the robustness of the pooled combined meta-analysis effects and between-study heterogeneity using the leave-one-out method. Moreover, we also performed *a priori* defined sensitivity analyses in the case that a number of small sample size studies would produce extreme effects on the pooled estimates. It is known that “heterogeneity-influencing” studies (small sample size studies favoring SA) may bias the pooled effects and that the induced heterogeneity may be exaggerated in the random-effect meta-analysis whereby the latter may become more biased than in the fixed-effect analysis.^48^ Thus, the aim is not to delineate the influencing studies or outlying studies but to examine whether the pooled estimates remain unchanged after performing a sensitivity analysis, which removed two or more heterogeneity-influencing studies from the CMA. If removal of the heterogeneity influencing cases does not alter the conclusions of the CMA, we can be confident that the pooled estimates are robust to influencing cases. In those cases, we will report the overall SMD as well as the SMD after excluding the heterogeneity-influencing studies.

Small study effects including publication bias was examined using the classical fail-safe N method, Kendall tau with continuity correction (using one-tailed p-values) and Egger’s regression intercept (using one tailed p-values). When Egger’s linear regression test suggests significant asymmetry, we use Duval and Tweedie’s trim-and-fill procedure to estimate the adjusted effect size after considering the effects of missing studies. Funnel plots, which display study precision in the y-axis and the SDM on the x-axis, are employed to detect small study effects or systemic heterogeneity by simultaneously displaying the observed studies and the imputed missing values.

## Results

### Search results

**Figure 2** shows the results of our search, selection and inclusion process. Out of 2.823 initial records identified via our search, 2.767 studies were removed. As such, 56 full text articles were assessed in the systematic review and for eligibility in the meta-analysis. Of those, another 5 were removed for reasons shown in **ESF Table 5** and, therefore, the meta-analysis was performed on 51 studies.^8–15, 17, 18, 38–43, 45, 47, 57–90^ and the systematic review on an additional 5 studies.^91–95^ Overall, the meta-analysis reports data from 29.093 participants, namely 4.945 SA+ patients and 24.148 controls with 8 studies reporting on 1.760 heathy controls, 23 studies reporting on 20.264 SA-patients, and 20 studies reporting on 2.124 SA-patients and healthy controls combined. Using these subgroups within the study as unit of analysis, we were able to compare 1.130 SA+ patients *versus* 2.778 healthy controls, and 3.815 SA+ patients *versus* 21.370 SA-patients. Seventeen studies reported data on a lifetime history of SA (1.205 SA+ patients *versus* 4.790 controls) and 30 on a current SA (3.412 SA+ patients *versus* 17.351 controls), whereas the other studies showed mixed data. Among SA+ patients, 1.124 suffered from MDD, 2.354 from affective disorders, 1.136 from mixed psychiatric disorders, and the others from schizophrenia, adjustment disorders, or other psychiatric disorders not including mood disorders. The age range of the participants was 12-83 years old. The meta-analysis included participants from all continents except Australasia, namely 1 study from Belgium, Germany, India, Iran, Japan, the Netherlands, Serbia, and Taiwan, 2 from Brazil, Canada, Croatia, France, and Poland, 3 from Iraq, 4 from China and Italy, 5 from South Korea, Sweden and Turkey, and 7 from the USA. Only 12 studies reported on the method of SA, and there are only 5 studies showing mean values of violent and non-violent SA. The other studies did not separate between violent and non-violent means. Only 20 studies specified the time of blood sampling. We included 45 case-control studies, 5 cohort studies, and 1 retrospective study. Twenty-three studies used serum to assay the biomarkers and 20 used plasma, whereas 5 reported the biomarkers were assayed in blood. The quality control scores are shown in ESF Table 3. The median quality control score was 5.94 (min= 1.56, max=9.69) and the median red point score was 17.75 (min=7.0, max=25.0).

**Figure 2.**
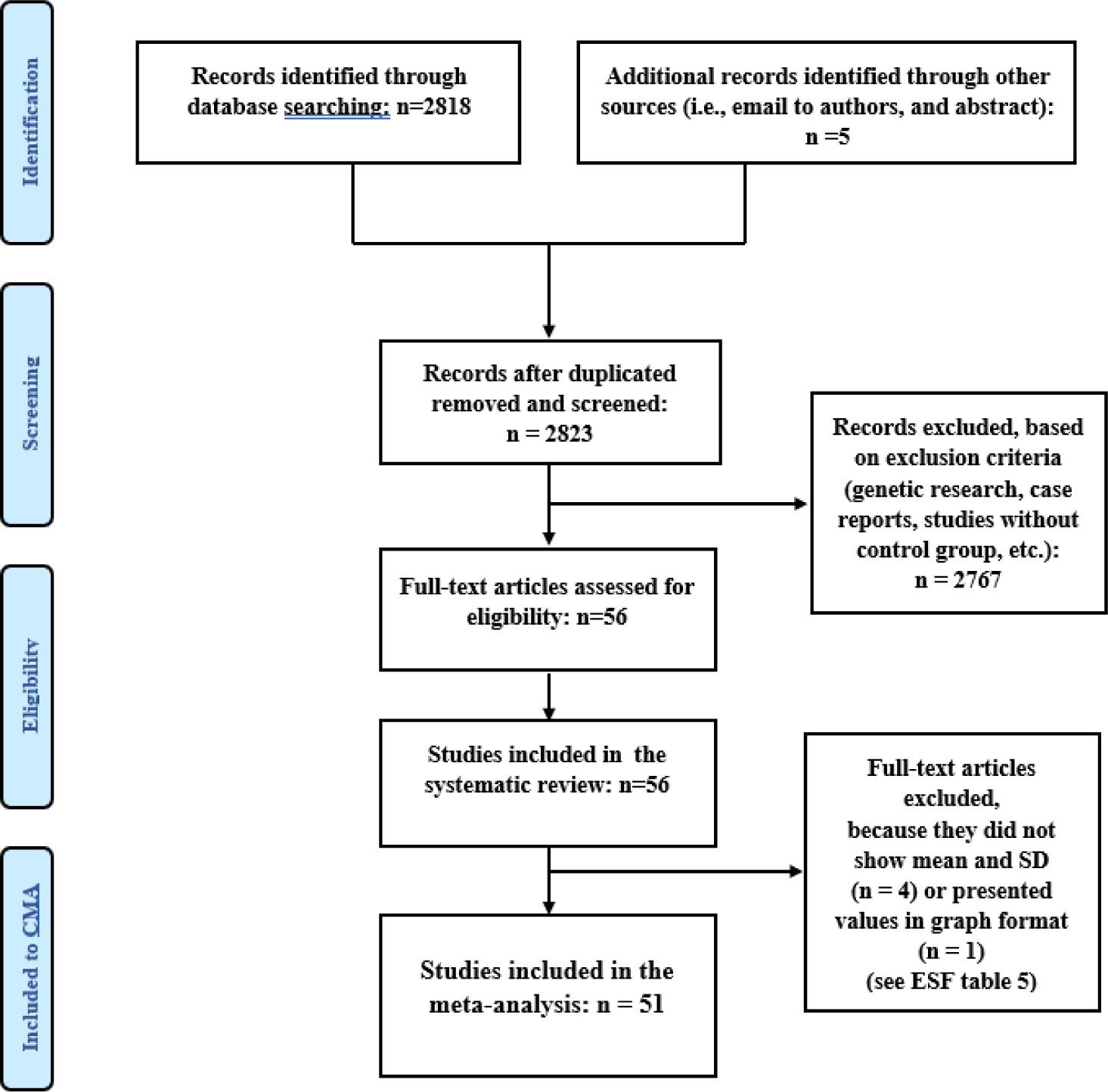
Prisma flow diagram.

### Main analysis of the primary outcome, IO&NS

**Table 1** shows the distributions of the SMD and confidence intervals with respect to the positive side of zero (favoring SA). We found that 25 studies showed confidence intervals that were entirely on the positive side of zero indicating statistically significant effects, whereas no confidence interval was entirely on the negative side of zero. The other (26 intervals) overlapped with zero with 20 studies showing SMD values that were greater than zero *versus 6* that were lower than zero. Moreover, ESF Table 5 displays that 4 out of the 5 studies^91–94^, which were not selected in the CMA, reported increased IO&NS biomarkers in SA+ patients as compare with SA- or controls. One study^95^ showed mixed results with increased TNF-α, but no changes in IL-1β, IL-6 or IL-2. **Figure 3** shows the forest plot of all IO&NS markers in SA versus controls. **Table 2** shows the results of the primary comparative random-effects meta-analysis performed on 51 studies and that the composite IO&NS levels were significantly higher in SA+ patients than in controls. Nevertheless, the heterogeneity indices suggested that considerable bias may be present and, therefore, we have performed subgroup analysis. When using the control subgroups as the unit of analysis, we found that the comparison of SA+ *versus* SA-yielded a SMD of 0.365, whereas the comparison of SA+ *versus* healthy controls yielded a large, pooled effect size of 0.826. Sensitivity analysis using the leave-one-out method did not alter the results of these meta-analysis (and any of the meta-analysis described below). Consequently, we performed a sensitivity analysis with heterogeneity-influencing studies removed. Table 2 shows that the SMD remained significant even after removing 6 heterogeneity-influencing studies favoring SA and that in the 20 studies, which compared SA+ with SA-patients, the heterogeneity was low (ᴦ^2^=0.006). No such reduction in heterogeneity could be found when using healthy controls as reference group. **Table 3** shows that there was some degree of publication bias with 7 missing studies on the right side of the funnel plot with an increased adjusted point estimate of 0.576 (95CI: 0.450; 0.701).

**Figure 3.**
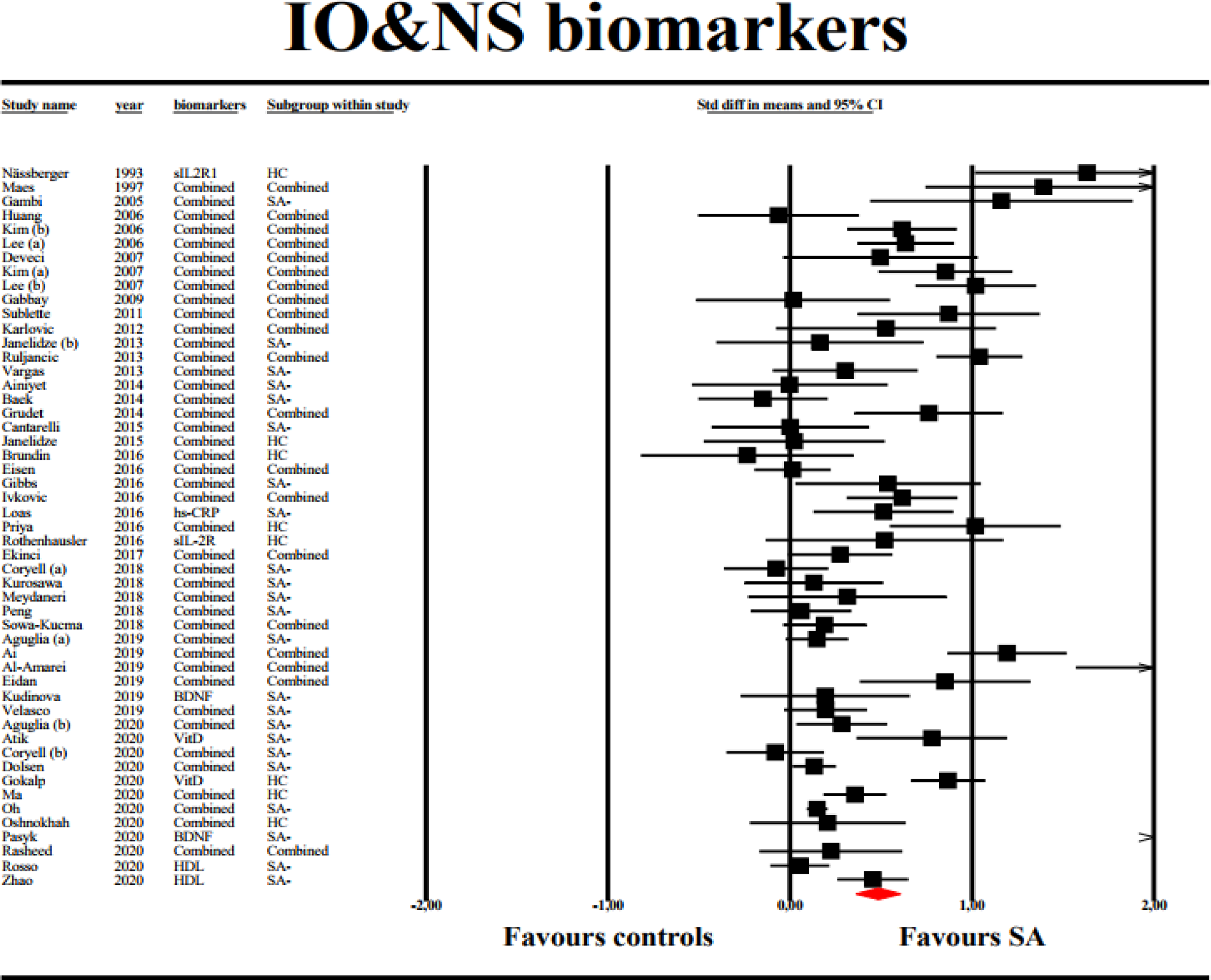
Forest plot with results of meta analysis performed on 51 studies reporting immune-inflammatory and oxidative & nitrosative stress (IO&NS) biomarkers.

**Table 2.** Results of meta-analysis performed on different outcome variables (immune and oxidative and nitrosative stress profiles, IO&NS, and subdomains)

| Outcome feature sets | n | Groups, subgroups | SMD | 95% CI | z | p | Q | df | p | I<sup>2</sup> (%) | τ<sup>2</sup> | T |
| --- | --- | --- | --- | --- | --- | --- | --- | --- | --- | --- | --- | --- |
| IO&NS | 51 | Overall | 0.485 | 0.363; 0.608 | 7.77 | <0.001 | 536.12 | 50 | <0.001 | 90.67 | 0.160 | 0.400 |
|  | 28 | SA+ vs HC | 0.826 | 0.607; 1.044 | 7.41 | <0.001 | 184.46 | 27 | <0.001 | 85.36 | 0.271 | 0.520 |
|  | 43 | SA+ vs SA- | 0.365 | 0.243; 0.487 | 5.86 | <0.001 | 314.18 | 42 | <0.001 | 86.63 | 0.119 | 0.344 |
|  | 35 | SA+ vs SA- | 0.162 | 0.101; 0.223 | 5.20 | <0.001 | 46.34 | 34 | 0.077 | 26.63 | 0.006 | 0.079 |
| IRS | 33 | Overall | 0.486 | 0.355; 0.616 | 7.28 | <0.001 | 243.85 | 32 | <0.001 | 86.88 | 0.107 | 0.327 |
|  | 15 | SA+ vs HC | 1.044 | 0.599; 1.489 | 4.60 | <0.001 | 140.28 | 14 | <0.001 | 90.02 | 0.674 | 0.821 |
|  | 25 | SA+ vs SA- | 0.290 | 0.183; 0.397 | 5.30 | 0.001 | 82.95 | 24 | <0.001 | 71.07 | 0.038 | 0.194 |
|  | 23 | SA+ vs SA- | 0.203 | 0.134; 0.273 | 5.70 | <0.001 | 31.64 | 22 | 0.084 | 30.47 | 0.006 | 0.079 |
| Inflammation | 22 | Overall | 0.466 | 0.311; 0.621 | 5.90 | <0.001 | 184.95 | 21 | <0.001 | 88.65 | 0.103 | 0.321 |
|  | 10 | SA+ vs HC | 1.109 | 0.505; 1.714 | 3.60 | 0.001 | 116.57 | 9 | 0.001 | 92.28 | 0.853 | 0.932 |
|  | 20 | SA+ vs SA- | 0.311 | 0.191; 0.432 | 5.05 | 0.001 | 77.52 | 19 | 0.001 | 75.49 | 0.042 | 0.205 |
|  | 7 | Lifetime | 0.180 | -0.012; 0.371 | 1.84 | 0.066 | 19.94 | 6 | 0.003 | 69.91 | 0.041 | 0.203 |
|  | 11 | Recent | 0.299 | 0.196; 0.402 | 5.57 | <0.001 | 15.24 | 10 | 0.124 | 34.39 | 0.008 | 0.089 |
| Neurotoxicity | 27 | Overall | 0.412 | 0.281; 0.544 | 6.13 | <0.001 | 170.25 | 26 | <0.001 | 84.73 | 0.085 | 0.291 |
|  | 14 | SA+ vs HC | 0.879 | 0.465; 1.293 | 4.16 | <0.001 | 113.67 | 13 | <0.001 | 88.56 | 0.530 | 0.729 |
|  | 23 | SA+ vs SA- | 0.315 | 0.198; 0.432 | 5.28 | <0.001 | 82.16 | 22 | <0.001 | 73.37 | 0.043 | 0.206 |
|  | 7 | 7 Lifetime | 0.115 | -0.032; 0.261 | 1.54 | 0.124 | 9.77 | 6 | 0.135 | 38.60 | 0.014 | 0.116 |
|  | 15 | 15 Recent | 0.318 | 0.260; 0.420 | 6.11 | <0.001 | 23.15 | 14 | 0.058 | 39.52 | 0.012 | 0.108 |
| ANTIOXPRO | 32 | Overall | 0.423 | 0.254; 0.593 | 4.90 | <0.001 | 456.47 | 31 | <0.001 | 93.21 | 0.204 | 0.452 |
|  | 16 | SA+ vs HC | 0.648 | 0.354; 0.941 | 4.33 | <0.001 | 133.89 | 15 | <0.001 | 88.80 | 0.295 | 0.543 |
|  | 28 | SA+ vs SA- | 0.341 | 0.167; 0.515 | 5.51 | <0.001 | 285.49 | 27 | <0.001 | 90.54 | 0.173 | 0.416 |

SMD: standardized mean difference, 95% CI: 95% confidence intervals
IO&NS: immune-inflammatory and oxidative and nitrosative stress
IRS: immune-inflammatory response system
ANTIOXPRO: protection via antioxidants and neurotrophic products.
We used the study as well as the prespecified subgroups as the units of analysis. Thus, patients with suicide attempts (SA+) were first (overall) compared with controls, that is healthy controls + patients without SA (SA-). Second, we performed subgroup analyses comparing the pooled effect size of IO&NS, IRS, inflammation, neurotoxicity and ANTIOXPRO profiles in a) SA+ versus SA- patients or healthy controls; and b) SA+ patients with a lifetime history (L) or recent (R) SA versus SA- patients and healthy controls.

**Table 3.**
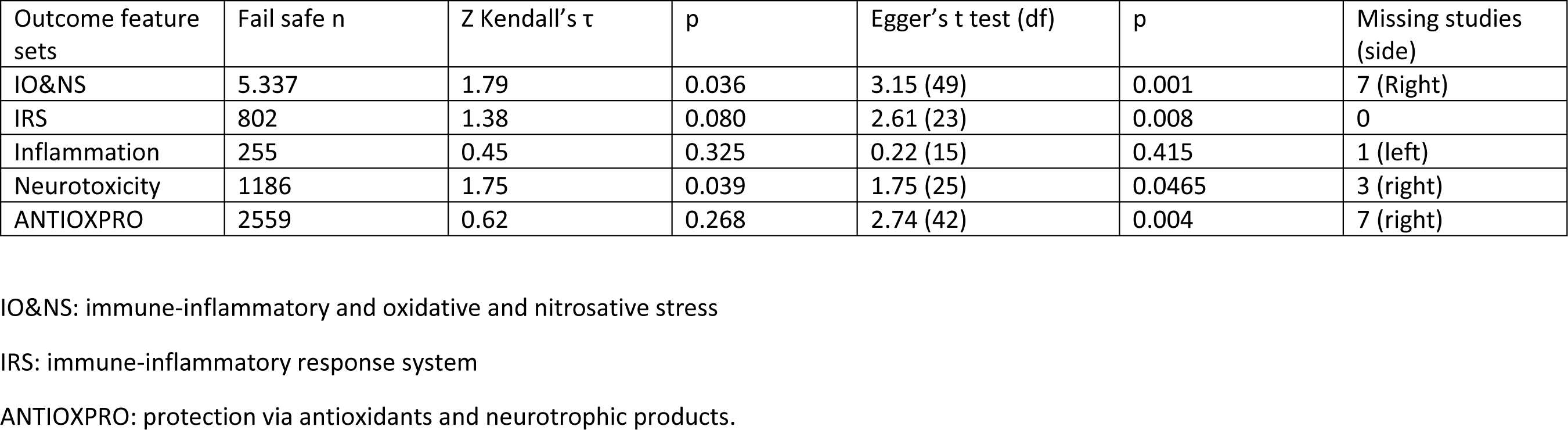
Results on publication bias.

### IRS profile

Table 1 shows the distribution of the confidence intervals obtained in the different studies regarding the positive side of zero. Eighteen studies showed confidence intervals with statistically significant effects, whereas no confidence intervals were entirely on the negative side of zero. Fifteen intervals overlapped with zero with 12 showing SMD values which were greater than zero and 3 SDM values lower than zero. ESF Table 5 shows that 4 out of the 5 studies^91–94^, which were excluded from the CMA, reported increased IRS biomarkers in SA+ patients as compared with SA- or controls and that one study^95^ showed mixed results with increased TNF-α, but no changes in IL-1β, IL-6 or IL-2. Table 2 shows that the composite IRS levels were significantly higher in SA+ patients than in controls, but the heterogeneity indices suggested some bias and, therefore, we have performed subgroup analysis (using the study as unit of analysis and the different control samples as subgroups). When using the control subgroups as the unit of analysis, we found that the comparison of SA+ *versus* SA-yielded a SMD of 0.290, whereas the comparison of SA+ *versus* healthy controls yielded a large, pooled effect size of 1.044. Consequently, we performed sensitivity analysis by removing 2 heterogeneity-influencing studies and found that when examining SA+ *versus* SA-, the SMD remained significant and that the heterogeneity indices were considerably decreased, and that the prediction interval was significant (0.0256; 0.3804). Table 3 shows that the impact of small study effects was minimal.

### Inflammation

Table 1 indicates that the 22 studies reporting on inflammation showed confidence intervals with statistically significant effects, whereas 0 confidence intervals were entirely on the negative side of zero and 9 intervals overlapped with zero. Of these, 6 showed SMD values that were greater than zero and 3 SMD values lower than zero. ESF Table 5 shows that 4 of the 5 studies^91–94^ that were excluded from the CMA, reported increased inflammatory biomarkers (CRP and TNF-α) in SA+ as compared with SA- or controls, although Conejero et al.^95^ found no significant changes in IL-1β and IL-6. **Figure 4** shows the forest plot of the inflammation profile and Table 2 shows that the composite inflammation score was significantly higher in SA+ patients than in controls, although there was considerable heterogeneity. When using the control subgroups as the unit of analysis, we found that the comparison of SA+ *versus* SA-yielded a SMD of 0.311 and the comparison of SA+ *versus* healthy controls a large, pooled effect size of 1.119. Using lifetime *versus* current SA improved heterogeneity and the latter was further improved by removing 4 heterogeneity-influencing studies favoring SA (see Table 2). Thus, when comparing recent SA+ with controls, the SMD (Table 2) and the prediction CI (0.0643; 0.5337) were both significant with an acceptable degree of heterogeneity (ᴦ^2^=0.008). However, when comparing a lifetime history of SA with controls, the SMD was not significant. We found a significant difference (p=0.002) among the effects at the current and lifetime levels. Table 3 shows that the impact of publication bias was minimal with 1 missing study on the left site of the funnel plot.

**Figure 4.**
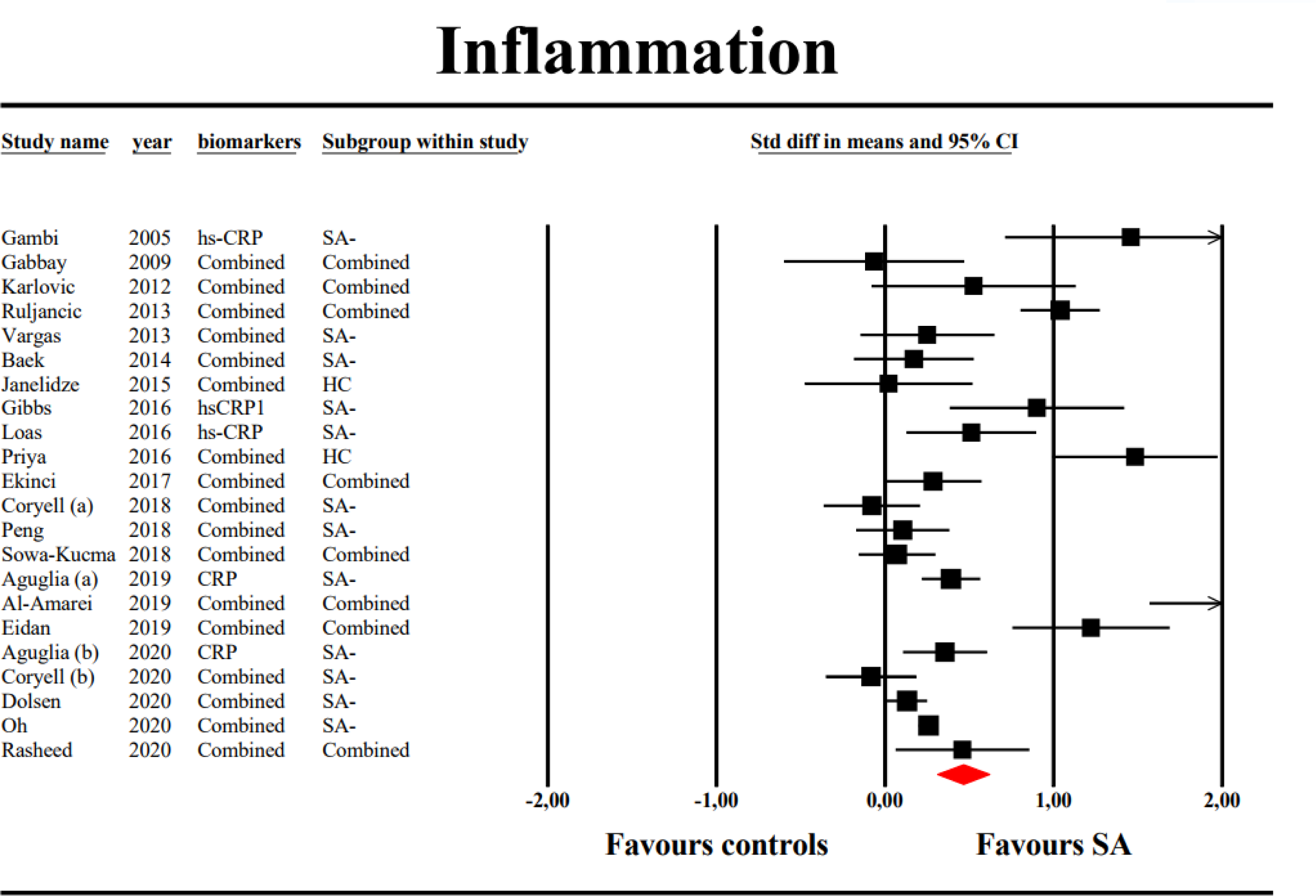
Forest plot with results of meta analysis performed on 22 studies reporting on inflammatory biomarkers.

### Neurotoxicity

Table 1 indicates that, out of the 27 studies reporting on neurotoxicity, 15 studies showed confidence intervals that were entirely on the positive side of zero, whereas there were no confidence intervals that were entirely on the negative side of zero. Twelve CI overlapped with zero, namely 8 showed SMD values that were greater than zero and 4 SMD values were lower than zero. Table 2 shows that the composite neurotoxicity score was significantly higher in SA+ patients than in controls, although there was some between-studies heterogeneity. When using the control subgroups as the unit of analysis, we found that the comparison of SA+ *versus* SA-yielded a SMD of 0.315 and the comparison of SA+ *versus* healthy controls a larger effect size of 0.879. Moreover, we performed a subgroup analysis based on the lifetime and current SA subgroups and performed a sensitivity analysis removing 4 heterogeneity-influencing studies favoring SA. When comparing recent SA+ with controls, we found that the SMD (Table 2) and the prediction CI (0.056; 0.580) were significant and that the impact of heterogeneity was low (ᴦ^2^=0.012). In contrast, when comparing a lifetime history of SA+ with controls, we found that the pooled effect size was not significant. Moreover, we found a significant difference (p=0.002) among the effects at the current and lifetime levels (p<0.001). Table 3 shows that the impact of small study effects was neglectable with 3 missing studies on the right side of the funnel plot.

### ANTIOXPRO profile

Table 1 shows that 14 studies reported confidence intervals that were entirely on the positive side of zero, whereas there were no confidence intervals that were entirely on the negative side of zero. Sixteen CI overlapped with zero, namely 11 showed SMD values that were greater than zero and 7 SMD values lower than zero. Table 2 shows that the ANTIOXPRO profile was significantly associated with SA, although heterogeneity was considerable. The effect size of the comparison SA+ versus healthy controls was larger than that of SA+ versus SA-. Performing subgroup analysis showed that the heterogeneity could not be reduced when comparing SA+ with SA-patients, or current and lifetime SA. Table 3 shows that there was some degree of publication bias with 7 missing studies on he right site of the funnel plot with an increased adjusted point estimate of 0.576 (95CI: 0.450; 0.701).

### Other subgroup analyses and meta-regression analyses

The associations of the primary outcome variable (IO&NS) with SA remained significant in patients with major depressive disorder (n=21, SDM: 0.622; 95%CI: 0.422; 0.822; z=6.11, p<0.001), mood disorders (n=12, SDM=0.155; 95%CI: 0.063; 0.247; z=3.31, p=0.001) and mixed study samples (n=13, SDM=0.581, 95%CI: 0.209; 0.953; z=3.06, p=0.002). The associations are also significant in serum and plasma samples and there are no significant differences between these media (p=0.770). Meta-regression did not reveal any effects of latitude, and both quality scores.

## Discussion

The major finding of this systematic review and meta-analysis is that SA is accompanied by signs of overall activation of IO&NS pathways, IRS, inflammation, and increased neurotoxicity, whereas the levels of antioxidants/neuroprotective factors may be decreased. To our knowledge, this is the first meta-analysis which combines different biomarkers into composites to delineate the associations between IO&NS, IRS, inflammation as well as neurotoxic profiles and SA. At first sight, it could be argued that we made a “fruit salad” by combining “apples and pears”, namely by combining different biomarkers. Nevertheless, the aim of our study was to compare IO&NS profiles between SA patients and controls. As such, we constructed various IO&NS composite scores by averaging the effects of the biomarkers in one and the same study and then averaging the effects all over the studies thereby examining IO&NS profiles rather than solitary markers.

As explained previously, it is much more relevant to combine IO&NS biomarkers into composite scores reflecting immune profiles rather than to examine the effects of solitary biomarkers.^6, 7, 30^ For example, in mood disorders the construction of O&NS composite scores revealed highly increased indicants of nitro-oxidative stress in major depression and bipolar disorder, whereas the differences in the solitary O&NS biomarkers were less evident.^7^ In schizophrenia, combining different neurotoxic IO&NS products into composites yielded more relevant results than considering the solitary biomarkers.^30^ It is now common practice to denote a small increase in plasma CRP concentrations in mood disorders as to indicate inflammation, although the increases in hsCRP are not in the range as detected in acute inflammatory disorders.^96^ These CRP results are, consequently, meta-analyzed and the small-moderate effect sizes obtained in the meta-analyses, which pooled small increases, are then interpreted to indicate evidence of “inflammation”.^37^ Nevertheless, in bipolar disorder, for example, a large part of the variance in hsCRP (up to 50%) is determined by the combined effects of body mass index, age, and early lifetime trauma casting doubts that a solitary assay of a small increase in hsCRP in mood disorders always reflect “inflammation”.^96^ However, in the current meta-analysis, we combined CRP data, at the case- and study level, with other inflammatory markers to obtain a synthetic score reflecting all available components of an inflammatory response, including CRP, other acute phase proteins such as albumin and fibrinogen and the erythrocyte sedimentation rate, together with the cytokines IL-1β, IL-6, TNF-α, and IL-8, which induce the acute phase response.^7^ As such, this study obtained a more robust synthetic index of “low-grade inflammation” as compared with a solitary hsCRP measurement, which may not be always useful. Using this synthetic score, we found significant low-grade inflammation in patients with SA as compared with controls (either healthy controls or SA-patients). These findings, therefore, consolidate the findings of a previous meta-analysis that suicidal behaviors are accompanied by increased levels of peripheral CRP indicating low-grade inflammation.^37^

Likewise, we also used synthetic scores which reflect IRS activation combining the inflammatory biomarkers with T helper (Th)-1 cytokines (IL-2 and IFN-γ), Th-1/Th-2 cell ratio (e.g. IL-2/IL-4; IFN-γ/IL-4), cytokine receptor levels (sIL-6R, sIL-2R, sIL-1RA, sTNFR60, sTNFR80), induction of the TRYCAT pathway (quinolinic acid levels), increased levels of NOx (produced during an immune response), and number of white blood cells, neutrophils, and the neutrophile/lymphocyte ratio (another marker of the immune response). These analyses showed IRS activation in SA+ as compared with controls. Such findings not only extend previous reports suggesting that immune-inflammatory cytokines (including IL-1) may be associated with suicidal behaviors^35, 36^, but also show that SA is accompanied by different indicants of IRS activation, combining low-grade inflammation with cell-mediated immune activation and its consequences (induction of TRYCAT pathway and NO production). In fact, our findings also extend previous reports that suicidal behaviors are associated with lowered 5-HT signaling^97^, because some immune-inflammatory cytokines, especially IFN-γ and IL-1, may induce indoleamine 2,3-dioxygenase (IDO) activity, which catabolizes tryptophan into kynurenines.^23^ Consequently, wider IRS activation (as observed in SA), may increase TRYCATs production (measured in this study) and decrease tryptophan levels leading to lowered 5-HT signaling in the brain.^23^

Another major finding is that the neurotoxicity synthetic score was significantly higher in SA patients as compared with controls. This neurotoxicity composite score comprises all neurotoxic IO&NS markers which are available in published original articles, namely cytotoxic cytokines such as IL-1β, IL-6 TNF-α, IL-2, and IFN-γ, the sIL-6R (which increases detrimental IL-6 trans-signaling), the chemokines IL-8, CCL-2, CCL-11, IP-10, and neurotoxic TRYCATs including kynurenine and quinolinic acid. ^6, 23, 30^ Another important part of the neurotoxicity score are increased levels of O&NS markers including NOx, LOOH, MDA, AOPP, and homocysteine.^23^

This meta-analysis also established that a composite score reflecting the neuroprotective potential, as indicated by BDNF and antioxidants such as albumin, vitamin D, TRAP, and TAC, is decreased in SA as compared with controls. However, this association is less robust in the meta-analysis as compared with the findings on IRS, inflammation and neurotoxicity because subgroup and sensitivity analysis did not reduce the heterogeneity. Nevertheless, the systematic review was highly indicative of lowered ANTIOXPRO and, therefore, these decreases in protective factors may predispose towards activated IRS and O&NS pathways and, therefore, IO&NS-associated neurotoxicity. Such findings indicate that SA is accompanied by increased neuro-immune and nitro-oxidative toxicity due to lipid peroxidation, aldehyde formation, chlorinative stress with protein oxidation, and maybe also increased nitrative and nitrosative stress, as a consequence of increased NO formation. All those phenomena are known to cause neurotoxicity with dysfunctions in grey and white matter functional plasticity in mood disorders and schizophrenia.^7, 30^ All in all, the results indicate that the same intertwined changes in CMI, low-grade inflammation, O&NS pathways and lowered antioxidant levels, which underpin the pathophysiology of mood disorders and schizophrenia, are associated with SA.

This meta-analysis found that the high heterogeneity in the different composite scores could be reduced through group analyses based on controls (SA- and healthy controls) and patients (current and lifetime SA). Furthermore, a sensitivity analysis, which removed heterogeneity-influencing studies that favor SA, further reduced heterogeneity. Firstly, the inflammation and neurotoxicity, but not IO&NS and IRS, pooled effect sizes and prediction intervals were significant when comparing current SA with controls, but not when comparing lifetime SA with controls. A current SA was defined as a SA within a time frame of maximal 1 month before blood sampling. Thus, it may be that SA is specifically associated with the effects of low-grade inflammation and enhanced neurotoxicity, and that a history of SA is associated with a sensitization of some IO&NS pathways. Previously, it was shown that recurrent depressive and manic episodes and suicidal behaviors cause a sensitization of immune (including IL-6, sIL-1RA, and sTNFRs) and O&NS (including MDA/TBARS) pathways and that staging of mood disorders is associated with increasing disabilities, O&NS (AOPP and MDA) and suicidal behaviors.^26^

Secondly, the effects sizes of the IO&NS, IRS, inflammation and neurotoxicity scores were large (from 0.776 to 1.119) when using healthy controls as reference group, but lower (around 0.300) when using SA-patients as reference group. The large effect sizes obtained with the normal group indicate that activated IO&NS pathways are strongly associated with SA in psychiatric disorders. The smaller effects sizes obtained when using SA- as reference group indicate that – within patients with major psychiatric disorders – further increases in IO&NS are associated with increased risk of SA. As such, activated IO&NS pathways with consequent neurotoxicity are not only associated with the major psychiatric disorders^7, 30^, but also with SA committed by those patients.

### Limitations

The results of this systemic review and meta-analysis should be discussed regarding its limitations. Firstly, the results of the meta-analysis showed high statistical heterogeneity in some IO&NS profiles as assessed with Q-values and the I^2^ metric. Nevertheless, there are considerable problems measuring heterogeneity with the Q test and the I^2^ metric suggesting that the latter may be of limited use to assess clinically relevant heterogeneity.^55^ Thus, Q increases with the number of studies in the analysis and with precision, whilst I^2^ increases with the precision and the number of patients included in the meta-analysis.^55^ Tau (τ) is a more accurate measurement of the true heterogeneity and, in contrast to the Q test and I^2^ metric, does not increase with number of patients or studies in the meta-analysis.^55^ When τ^2^ was imprecise, we performed random-effects subgroup meta-analysis considering a priori defined patient and control subgroups and found that the latter considerable decreased heterogeneity, especially when combined with sensitivity analysis, which removed a set of heterogeneity-influencing studies. Most importantly, the latter sensitivity analyses showed that the effect sizes of SA+ *versus* SA- and current SA *versus* controls remained significant and even showed significant prediction intervals. This indicates that the results of these meta-analyses are robust to heterogeneity.

Secondly, in the 51 included studies, there were only few data on suicide methods (violent *versus* non-violent) and, therefore, we were unable to perform group analyses. This is another source of statistical heterogeneity because violent suicide is more strongly associated with different immune and metabolic biomarkers than non-violent suicide.^33^ Thirdly, we were unable to construct other important immune profiles including the T regulatory (Treg), Th-2 and CIRS profiles. Thus, few papers measured IL-4^12, 16^ or TGF-β1^17^ and, therefore, these studies could not be included in the CMA. In addition, other cytokines of these profiles are completely missing, including the most important negative-immune regulatory cytokine, IL-10. Moreover, there are no Treg and T effector CD expression data in SA, although such flow cytometric measurements are strongly associated with staging of mood disorders, which, in turn is strongly related with suicidal behaviors.^98^

Fourthly, although our aggregate indices comprise many important biomarkers, others are lacking in the reviewed literature. Future research in SA should include more biomarkers not only to assess Treg, Th-2, and CIRS profiles, but also to enrich the profiles assessed in the current meta-analysis (see ESF, Table 1, column: what is missing in SA research). For example, the synthetic index of inflammations should incorporate more positive (especially haptoglobin) and negative (especially transferrin and zinc) acute phase reactants.^22^ The O&NS profile should include markers of oxidative damage to DNA/RNA and mitochondria, as well as key antioxidant markers including paraoxonase 1, glutathione, and coenzyme Q10.^24, 29^ Future research on SA in mood disorders should incorporate the effects of staging of illness and its related biomarkers including O&NS.^98^

### Conclusions

SA are associated with IO&NS, IRS activation, inflammation, increased neurotoxicity and lowered neuroprotection. A current SA is strongly associated with inflammation and increased neurotoxicity, and patients with a lifetime history of SA show a sensitized IRS system. The strong associations between SA and increased neurotoxicity may be explained by the combined neurotoxic effects of pro-inflammatory cytokines, TRYCAT pathway activation, and O&NS including increased NO production. Lowered neuroprotection due to lowered antioxidant and neurotrophic defenses may further aggravate those neurotoxic effects. This meta-analysis discovered new biomarkers and therapeutic targets to treat individuals with suicidal behaviors, which accompany psychiatric disorders. Figure 1 shows the different neurotoxic pathways that are involved in the major psychiatric disorders and SA as well.

## Ethical approval

None.

## Author contributions

All authors contributed to the writing up of the paper. The work was designed by MM and AV. Data were collected by AV and KJ. Statistical analyses were performed by MM. All authors revised and approved the final draft.

## Data availability

The dataset generated during and/or analyzed during the current study will be available from MM upon reasonable request and once the dataset has been fully exploited by the authors.

## Supplementary files

**ESF1 Table 1.**
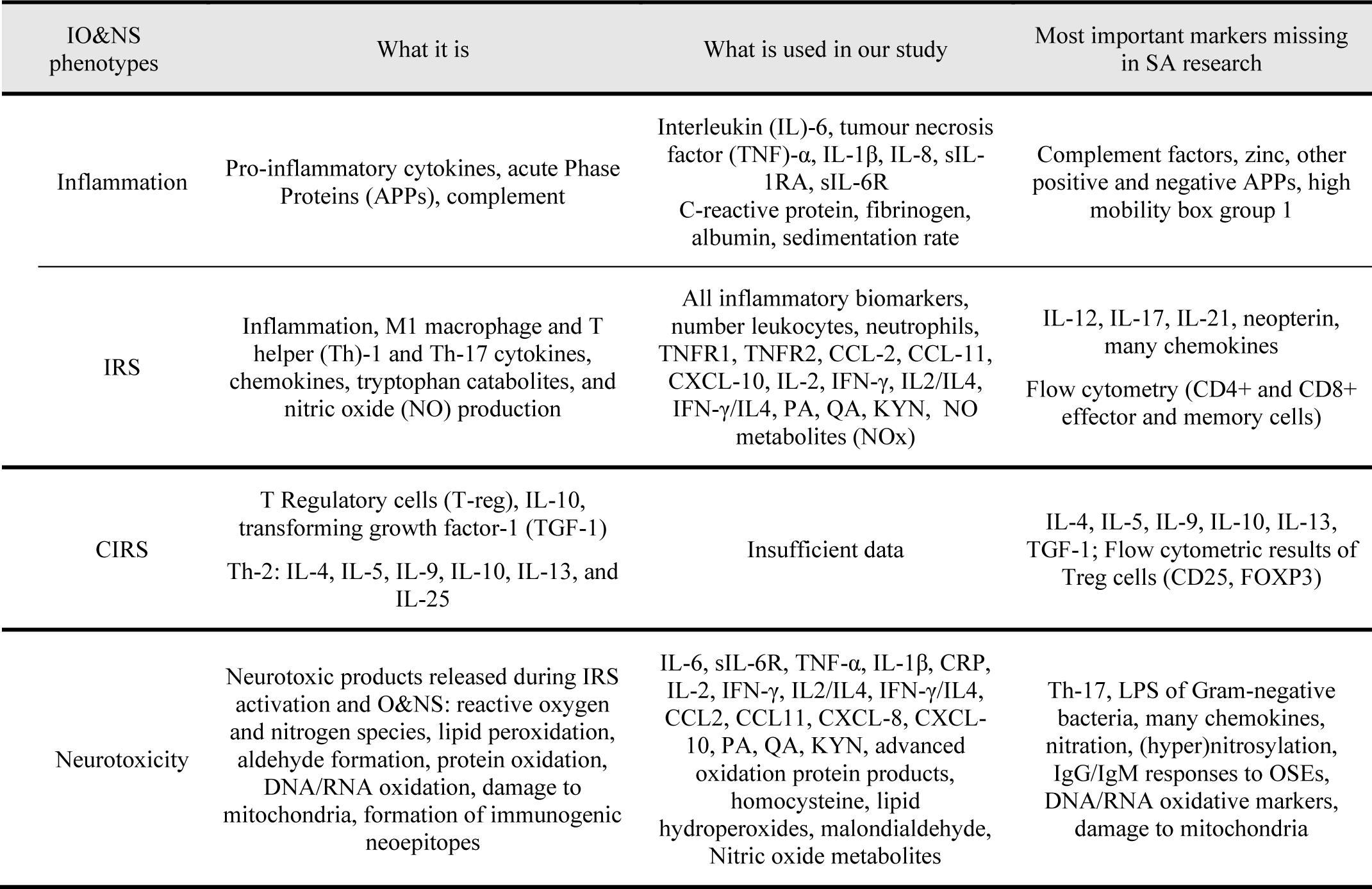
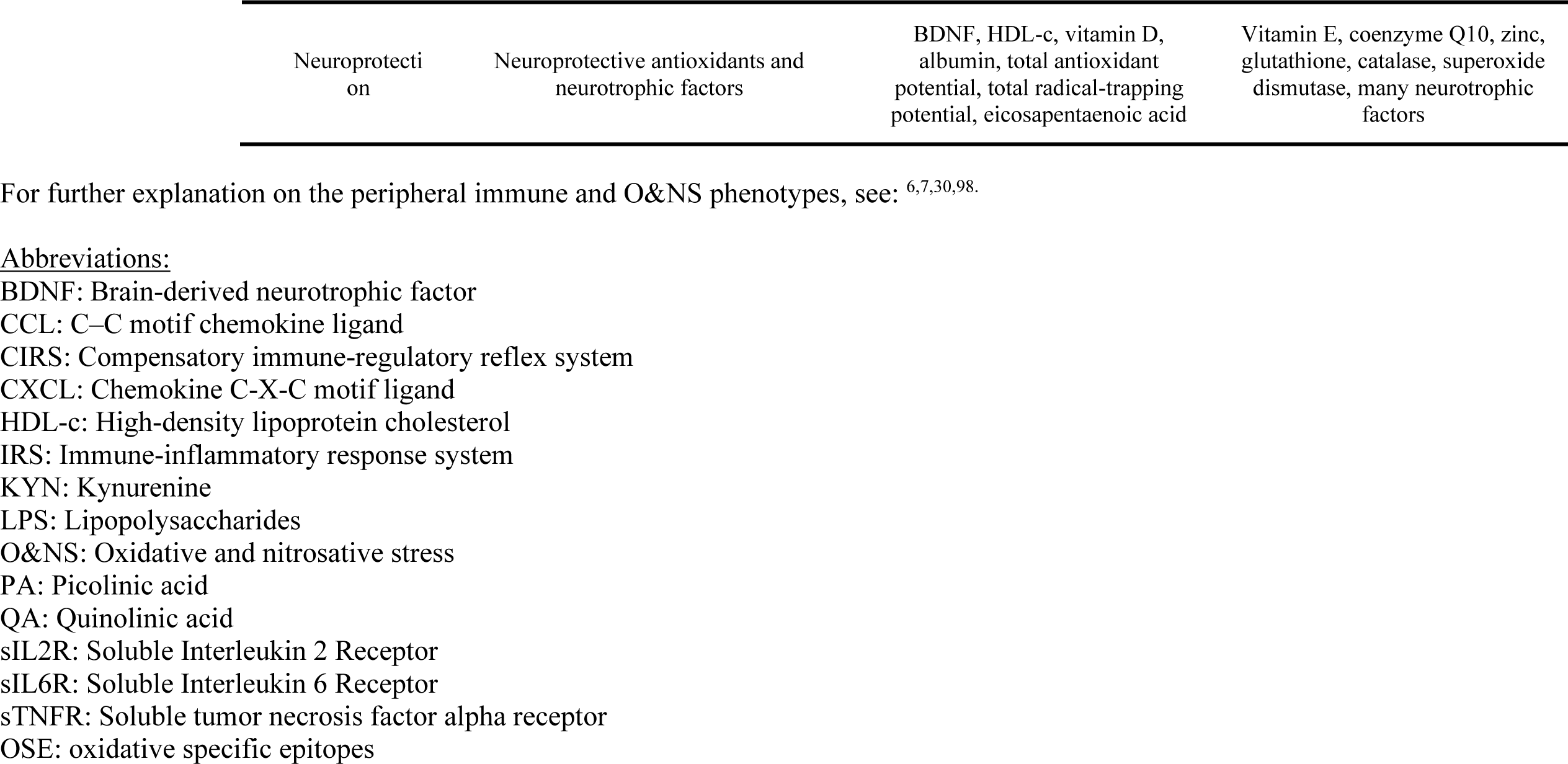
Peripheral immune-inflammatory and oxidative and nitrosative stress (IO&NS) phenotypes in suicide attempts (SA).

**ESF Table 2.** Specific search for each database.

|  |
| --- |
| **PubMed/Medline** |
| ((inflamm* OR immun*) AND (cytokine OR chemokine OR il-6 OR il-1 OR interleukin OR (c-reactive protein OR CRP) OR (Tumor necrosis factor OR tnf) OR (interferon OR ifn) OR (Transforming growth factor OR tgf) OR Tryptophan AND (suicid*)) **1026** |
| **PubMed/Medline** |
| ((Oxidative stress OR Antioxidants) AND (Zinc) OR (Vitamin C) OR (Albumin) OR (Nitric oxide metabolites) OR (Lipid hydroperoxides) OR (Omega 3) OR (Coenzyme Q10) OR (DHA) AND (suicid*)) **383** |
| **Google Scholar** |
| Suicide AND [inflamm*] OR [immun*] AND [oxidative stress biomarkers] AND antioxidants OR [cytokine] OR chemokine] OR [il-6 OR il-1 OR interleukin] OR [(c-reactive protein) OR CRP] OR [(Tumor necrosis factor) OR tnf] OR [interferon OR ifn] OR (Transforming) **525** |
| **WEB OF SCIENCE** |
| (suicide) AND TOPIC: (inflammatory biomarkers) **35** |
| **WEB OF SCIENCE** |
| (suicide) AND ALL FIELDS: (oxidative stress biomarkers) **17** |
| **WEB OF SCIENCE** |
| TOPIC: (suicide) AND ALLFIELDS: (("inflamm*" OR "immun*") AND ("cytokine" OR "chemokine" OR "il-6" OR "il-1" OR "interleukin" OR "c-reactive protein" |

**ESF Table 3.**
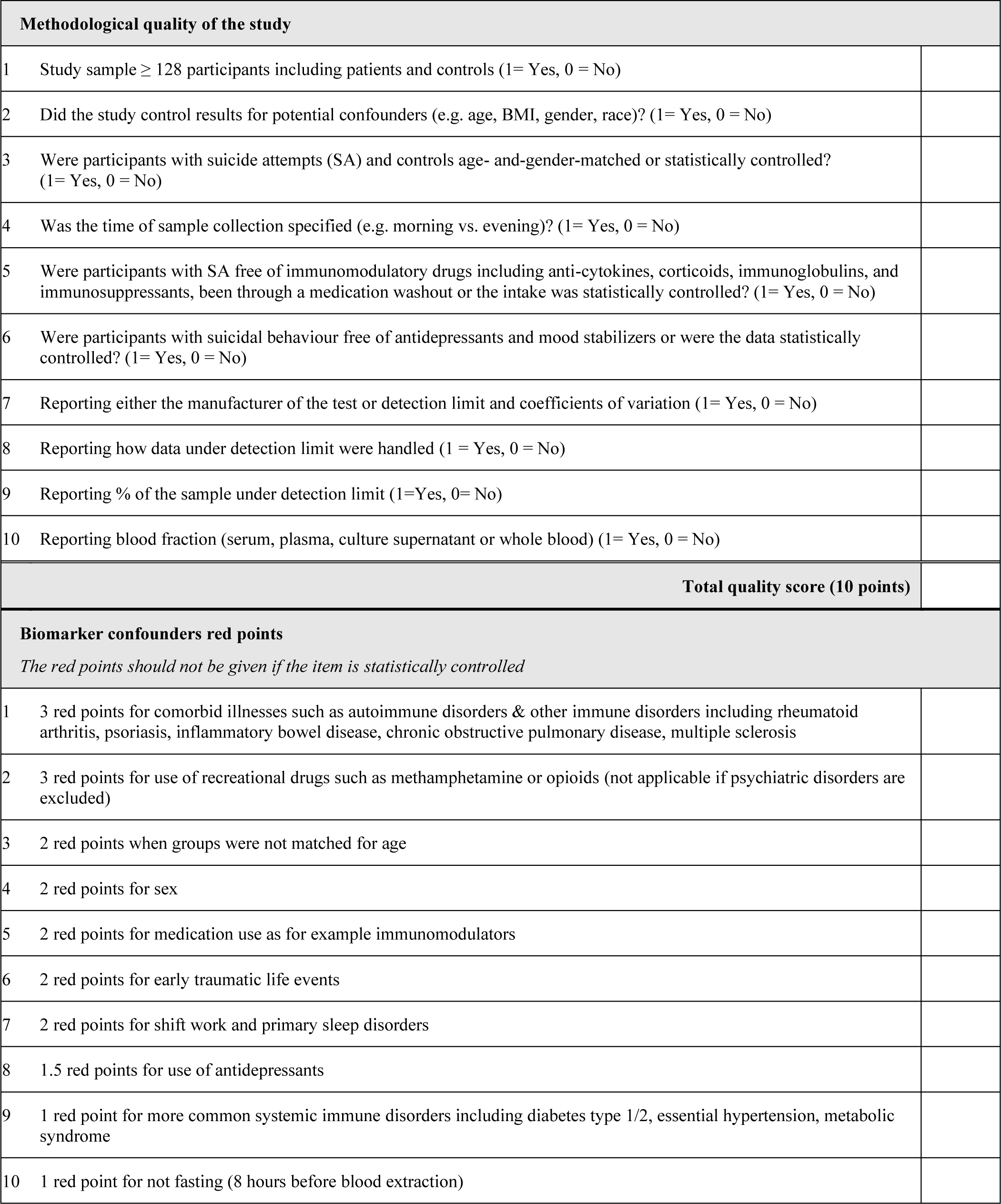
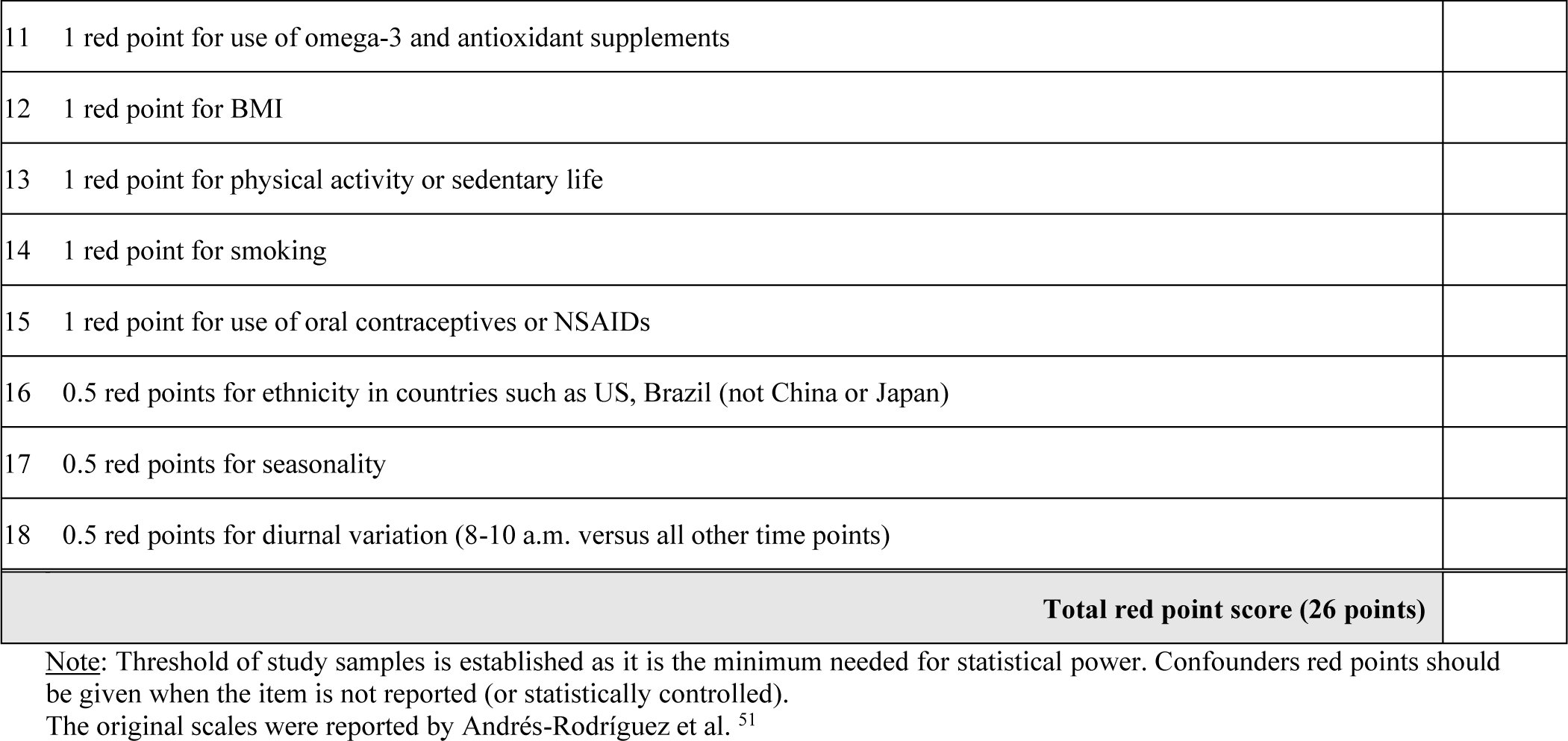
Immune cofounder’s scale (ICS) applied from Andrés-Rodríguez, et al., 2019^51^.

| **Methodological quality of the study** |  |
| --- | --- |
| 1 | Study sample $\geq 128$ participants including patients and controls (1= Yes, 0 = No) |
| 2 | Did the study control results for potential confounders (e.g. age, BMI, gender, race)? (1= Yes, 0 = No) |
| 3 | Were participants with suicide attempts (SA) and controls age- and-gender-matched or statistically controlled? (1= Yes, 0 = No) |
| 4 | Was the time of sample collection specified (e.g. morning vs. evening)? (1= Yes, 0 = No) |
| 5 | Were participants with SA free of immunomodulatory drugs including anti-cytokines, corticoids, immunoglobulins, and immunosuppressants, been through a medication washout or the intake was statistically controlled? (1= Yes, 0 = No) |
| 6 | Were participants with suicidal behaviour free of antidepressants and mood stabilizers or were the data statistically controlled? (1= Yes, 0 = No) |
| 7 | Reporting either the manufacturer of the test or detection limit and coefficients of variation (1= Yes, 0 = No) |
| 8 | Reporting how data under detection limit were handled (1 = Yes, 0 = No) |
| 9 | Reporting % of the sample under detection limit (1=Yes, 0= No) |
| 10 | Reporting blood fraction (serum, plasma, culture supernatant or whole blood) (1= Yes, 0 = No) |
| **Total quality score (10 points)** |  |
| **Biomarker confounders red points** |  |
| *The red points should not be given if the item is statistically controlled* |  |
| 1 | 3 red points for comorbid illnesses such as autoimmune disorders & other immune disorders including rheumatoid arthritis, psoriasis, inflammatory bowel disease, chronic obstructive pulmonary disease, multiple sclerosis |
| 2 | 3 red points for use of recreational drugs such as methamphetamine or opioids (not applicable if psychiatric disorders are excluded) |
| 3 | 2 red points when groups were not matched for age |
| 4 | 2 red points for sex |
| 5 | 2 red points for medication use as for example immunomodulators |
| 6 | 2 red points for early traumatic life events |
| 7 | 2 red points for shift work and primary sleep disorders |
| 8 | 1.5 red points for use of antidepressants |
| 9 | 1 red point for more common systemic immune disorders including diabetes type 1/2, essential hypertension, metabolic syndrome |
| 10 | 1 red point for not fasting (8 hours before blood extraction) |
| 11 | 1 red point for use of omega-3 and antioxidant supplements |
| 12 | 1 red point for BMI |
| 13 | 1 red point for physical activity or sedentary life |
| 14 | 1 red point for smoking |
| 15 | 1 red point for use of oral contraceptives or NSAIDs |
| 16 | 0.5 red points for ethnicity in countries such as US, Brazil (not China or Japan) |
| 17 | 0.5 red points for seasonality |
| 18 | 0.5 red points for diurnal variation (8-10 a.m. versus all other time points) |
| **Total red point score (26 points)** |  |

Note: Threshold of study samples is established as it is the minimum needed for statistical power. Confounders red points should be given when the item is not reported (or statistically controlled). The original scales were reported by Andrés-Rodríguez et al.<sup>51</sup>

